# Gender Trends in Youth Tobacco and Nicotine Use in Georgia Across GYTS Rounds, 2014–2023

**DOI:** 10.1101/2025.09.09.25335320

**Authors:** Kakha Gvinianidze, George Bakhturidze, Tamar Abuladze, Ana Dekanosidze, Lela Sturua, Veriko Gegenava, Yelena Tarasenko, Angela Ciobanu

## Abstract

**Background:** Adolescent tobacco and nicotine product (TNP) use remains a public health concern, with emerging gender convergence and rising e-cigarette use among females. We examined gender-specific trends in Georgian adolescents alongside national tobacco control policies and social context.

**Methods:** We analyzed nationally representative Global Youth Tobacco Survey data from Georgia (2014, 2017, and 2023) for youth aged 11-17 to estimate prevalence and patterns of current TNP use in the context of tobacco-related advertising and promotion, secondhand smoke, and national policy, via a policy review. Adjusted prevalence estimates were derived using post-estimation predictive margins after fitting multinomial logistic regression models.

**Results:** From 2014 to 2023, current TNP use declined among boys but increased among girls, particularly in exclusive e-cigarette use. The percentages reporting no current TNP use increased among boys from 77.4% to 85.6% (p < 0.01), but declined among girls from 92.1% to 88.1% (p = 0.04). By 2023, exclusive e-cigarette use was more prevalent among girls than boys. Across sexes, exposure to pro-tobacco advertising and promotion and secondhand smoke declined.

Exposure to anti-tobacco promotion peaked in 2017 and receded by 2023. The policy review documented strengthened measures, alongside gaps in oversight of digital marketing and newer products, including e-cigarettes.

**Conclusions:** Georgia’s tobacco control policies have contributed to reduced TNP use among boys and a less permissive social environment. Rising e-cigarette use among girls is concerning. Policies should close regulatory gaps, enhance enforcement, and address evolving product and marketing developments to prevent TNP uptake and protect adolescents from tobacco-related harms.

**SUMMARY BOX:** *What is already known on this topic:* Adolescent tobacco use patterns are shifting globally, with narrowing gender gaps and rising use of new products like e-cigarettes, especially among girls. In Georgia, the impact of recent tobacco control reforms targeting youth tobacco and nicotine product use by gender had not been fully assessed.

*What this study adds:* Between 2014 and 2023, TNP use significantly declined among boys in Georgia but rose in exclusive e-cigarette use among girls. Exposure to tobacco advertising and promotion and secondhand smoke decreased, yet gaps in anti-tobacco promotion and regulation of novel products persist.

*How this study might affect research, practice or policy:* Findings from this study reinforce WHO’s call for gender-responsive tobacco control policies. Regulatory measures must address how product design and marketing tactics exploit gendered social norms and youth vulnerabilities. Comprehensive, gender-responsive strategies that strengthen anti-tobacco promotion and close regulatory gaps are essential to counter these influences and protect all adolescents from targeted industry tactics.

## INTRODUCTION

The World Health Organization (WHO) Framework Convention on Tobacco Control (FCTC) and most national tobacco control laws aim to ensure that all population groups benefit from protective measures.^1^ However, tobacco and nicotine industries continue to exploit gender-specific values, aspirations, and social norms to tailor product design and marketing strategies.^2^ Historically, when uptake slowed in one gender, companies shifted tactics to grow use among the other, sustaining high overall prevalence.^3-8^ Evidence from the early 2000s confirmed this pattern, with the Global Youth Tobacco Survey (GYTS) results showing convergence in smoking rates between boys and girls across multiple WHO regions, and in some countries, girls even surpassing boys.^5^ This gender catch-up trend underscores the need for timely, gender-responsive prevention efforts, particularly as a similar pattern may now be emerging with new tobacco and nicotine products (TNPs).

Yet, nearly two decades after the WHO Kobe Declaration, gender-responsive action in tobacco control remains limited.^7,8^ A 2018 WHO policy brief noted that while industry has long manipulated gender norms to expand its market, public health responses often remain gender-blind, lacking targeted interventions or meaningful evaluation of gendered effects.^8^

These evolving patterns underscore the urgency of preventive actions. Tobacco use causes over 7 million deaths annually, including approximately 1.6 million from secondhand smoke exposure.^9^ Among adolescents aged 13–15 years, at least 37 million (9.7%) use some form of tobacco, with about 4 million (10.8%) of them in the WHO European Region.^10^

Gender-responsive marketing strategies have continued to evolve alongside the rapid expansion of novel tobacco and nicotine products (TNPs), including electronic cigarettes (e-cigarettes), heated tobacco products (HTPs), and nicotine pouches. Features such as flavorings, sleek designs, and claims of reduced harm are often used to target adolescents by minimizing perceived risks and enhancing product appeal.^11-15^ Regulatory responses have often lagged behind these developments, leaving policy gaps and blind spots that facilitate youth access and reinforce misleading perceptions of reduced harm.^8^

Despite Georgia’s adoption of the comprehensive Tobacco Control Law in 2017, regulating TNPs remains complex.^16^ As in many countries, Georgia legislation tends to react rather than anticipate developments in the tobacco industry, which continuously introduces new product types product design characteristics, and marketing strategies. For example, nicotine-free alternatives, nicotine analogues, flavourings, and certain design and packaging formats are not always fully addressed by existing regulations.^16^ Variations in labeling, health warnings, and taxation across product categories may further reduce the overall effectiveness of control measures, particularly among young people. Meanwhile, alongside its well-documented health and social consequences, tobacco use continues to impose a substantial economic burden on Georgia, costing an estimated USD 341 million annually, or roughly 2.4% of the country’s GDP.^17^

Understanding gender-specific patterns in tobacco use is critical for ensuring that policy measures remain responsive and effective while upholding principles of fairness under law.^7,8^ This study reviewed the state of tobacco legislation in Georgia and examined social context and trends in youth TNP use by gender in 2014, 2017, and 2023. By doing so, we aimed to identify opportunities for proactive, equitable, and evidence-informed approaches to address emerging gender-specific patterns in youth tobacco use.

## METHODOLOGY

This observational study used cross-sectional data from three rounds of the GYTS conducted in Georgia in 2014, 2017, and 2023 by the National Center for Disease Control and Public Health (NCDC) under the Ministry of Health. The GYTS is a nationally representative, school-based survey of students aged 13–15 years that monitors product use, awareness of health risks, exposure to tobacco marketing, and support for tobacco control policies among school-aged adolescents.^18^

The GYTS uses a standardized two-stage cluster sampling design: schools are selected with a probability proportional to enrollment size, and grades 7-10 are randomly chosen; all students in these classes are eligible to participate. Although the core target population is students aged 13– 15 years, other students present on the survey day may also participate. We included all respondents regardless of age to maximize statistical power and ensure comparability across survey rounds. Data were collected anonymously using a self-administered questionnaire completed in classrooms on scannable answer sheets.

In 2014, there were 1,379 students-participants, including 962 aged 13–15 years, with an overall response rate of 75.4%. In 2017, there were 1,345 respondents (954 aged 13–15 years), and the overall response rate was 78.7%. The 2023 round included 2,569 respondents, of whom 1,845 were aged 13–15 years, with a response rate of 79.1%.^18^

The survey instrument included a standardized core set of questions and optional items tailored to national tobacco control priorities. We focused on four product types measured consistently across all three rounds: (1) cigarettes (manufactured or hand-rolled); (2) other smoked tobacco (e.g., pipes, cigars, mini-cigars/cigarillos, and shisha); (3) smokeless tobacco (e.g., chewing tobacco, snuff, snus); and (4) e-cigarettes. Consistent with the WHO definition of current use (or ‘past-month use’), we operationalized current cigarette and e-cigarette use as having used the product one or more days in the past 30 days. Use of other smoked and smokeless tobacco products was assessed by yes/no questions regarding use in the past month. A constructed composite variable captured the total number of TNP types used (range: 0–4). Participants missing data for any product type were coded as missing for this count to avoid underestimation (excluded: 217/1,379 in 2014; 173/1,345 in 2017; and 455/2,569 in 2023).

We created three composite indices to assess tobacco-related social influences: (1) anti-tobacco promotion exposure (protective factors), (2) pro-tobacco advertising and promotion exposure (risk factors), and (3) exposure to secondhand smoke, based on five, four, and six items, respectively. Each index was constructed by summing rescaled items (range: 0-1) from self-reported survey questions on educational messaging, advertising and promotion, interpersonal communication, or second-hand exposure. Given the conceptual diversity of the items, spanning various settings, sources, and tobacco-related behaviors, the indices were intended as additive exposure composites rather than internally consistent scales.^19^ The net tobacco promotion exposure index was calculated as the difference between anti- and pro-tobacco advertising and promotion exposure indices. Lower or negative scores indicate stronger exposure to anti-tobacco promotion, while higher (positive) scores reflect greater exposure to pro-tobacco advertising or promotion.

As part of the core questionnaire, students also reported their age and sex. In all three rounds, the optional section included a question on the range of students’ pocket money. We included this variable in our analyses as a proxy for socioeconomic status due to its relevance and modifiable nature, particularly in relation to youth purchasing behavior.^20^

### Statistical Analysis

Descriptive statistics included weighted percentages for categorical variables (age, sex, and pocket money) and weighted means for continuous variables (age and the four indices), each with 95% confidence intervals (CIs) by survey round. Crude and adjusted prevalence estimates for current use of none, one, or multiple TNPs were calculated overall, by sex, and by survey year. Adjusted estimates were derived using average marginal predictions from multinomial logistic regression models with age, sex, and pocket money as covariates.

Sex-based patterns observed in previous surveys and programmatic work of public health professionals in Georgia, motivated the conduct of stratified analysis, and these differences were subsequently confirmed in our data. When analyses were stratified by sex, prevalence estimates were adjusted for age and pocket money only. For the analyses of sex differences in current exclusive use of cigarettes, e-cigarettes, or other TNPs, stratified models were not feasible due to limited sample size. Predictive margins were calculated from the unadjusted and adjusted multinomial logistic regression models with the interaction between sex and year in effort to provide more stable estimates of outcome probabilities across sex and year.

To account for the complex survey design in the multivariable analyses, data from the three rounds were combined into a single dataset. Sampling weights were divided by the number of rounds to avoid inflating the effective sample size and to ensure appropriate variance estimation. Primary sampling units (PSUs) and strata were recoded to create unique identifiers across survey rounds, preserving PSU independence within each round. All analyses were restricted to respondents with complete data for the dependent and independent variables.

As a sensitivity check, we compared the socio-demographic profiles (age, sex, grade, pocket money) of students with and without complete TNP use data for each round. No meaningful differences were observed, suggesting minimal bias due to missingness.

All statistical analyses accounted for the complex survey design and were conducted using Stata version 17. A two-tailed alpha of 0.05 was used for statistical significance.

In addition to quantitative analyses, we conducted a descriptive review of tobacco control policies implemented in Georgia between 2014 and 2023. This review drew on publicly available legislative documents, tax codes, and regulatory updates to summarize key national measures related to taxation, packaging, advertising, retail restrictions, and product regulation.^16, 21-25^ The review focused on policy changes relevant to youth access and exposure, particularly those affecting conventional cigarettes, roll-your-own tobacco, smokeless tobacco, and electronic nicotine delivery products. In accordance with Georgian legislation, e-cigarettes, and other nicotine products are classified as tobacco products.^16^

The GYTS datasets are publicly available and de-identified. Therefore, this analysis did not constitute human subjects research and was exempt from institutional ethics review.

## RESULTS

### Study Population

Across the three survey rounds, most students were aged 13–15 years, with balanced gender representation and stable patterns of pocket money. Socio-demographic characteristics were broadly consistent across survey rounds (Table 1).

**Table 1.** Socio-Demographic Characteristics of Students in Grades 7–10 in Georgia.

|  | 2014 |  | 2017 |  | 2023 |  |
| --- | --- | --- | --- | --- | --- | --- |
|  | n | Weighted %<br>(95% CI) | n | Weighted %<br>(95% CI) | n | Weighted %<br>(95% CI) |
| **Age** |  |  |  |  |  |  |
| Mean age in years | 1,372 | 14.<br>(13.8 – 14.1) | 1,342 | 14.2<br>(14.0 – 14.4) | 2,563 | 13.4<br>(13.4 – 13.5) |
| Missing | 7 | – | 3 | – | 6 | – |
| **Gender** |  |  |  |  |  |  |
| Boys | 670 | 52.1<br>(46.3 – 57.8) | 659 | 50.8<br>(47.4 – 54.1) | 1220 | 51.8<br>(50.0 – 53.6) |
| Girls | 684 | 47.9<br>(42.2 – 53.7) | 673 | 49.2<br>(45.9 – 52.6) | 1319 | 48.2<br>(46.4 – 50.0) |
| Missing | 25 | – | 13 | – | 30 | – |
| **Pocket money** |  |  |  |  |  |  |
| Below median to none | 933 | 64.7<br>(56.2 – 72.4) | 887 | 65.3<br>(58.8 – 71.3) | 1,640 | 63.9<br>(59.1 – 68.5) |
| Median | 111 | 8.2<br>(6.7 – 10.0) | 95 | 7.0<br>(5.4 – 8.9) | 323 | 12.8<br>(11.2 – 14.6) |
| Above median | 329 | 27.1<br>(20.3 – 35.1) | 346 | 27.8<br>(22.4 – 33.9) | 571 | 23.2<br>(19.8 – 27.2) |
| Missing | 6 | – | 17 | – | 35 | – |

*Notes:* n–unweighted number of observations. Pocket money – money to spend on yourself in an average week.

**Table 2.** Summary of National Regulations in Georgia on Different Types of Tobacco Products^1^.

| Products | Year | Sales ban | Plain packaging | Health warnings (pictorial and textual health warning) | Flavor ban | Point of sale display ban (Internal & external visibility) | Advertising ban | Smoking ban in public places<sup>[1]</sup> | Age limit | Taxes* (GEL) |
| --- | --- | --- | --- | --- | --- | --- | --- | --- | --- | --- |
| Conventional cigarette | 2014 | – | – | Pictorial warnings were not mandatory | – | Only in those sections, where children's clothing and toys are sold | – | Partially | ? | Filtered – 0.9 GEL<br>Unfiltered – 0.25 GEL |
|  | 2017 | – | – | ? | – | Only in those sections, where children's clothing and toys are sold | ? | Partially | 18 | Filtered – 1.7 GEL<br>Unfiltered – 0.6 GEL |
|  | 2019 | – | – | ? | – | Only external visibility | ? | ? | 18 | 1.7 GEL |
|  | 2023 | – | Was adopted but not in force | ? | – | ? | ? | ? | 18 | 1.7 GEL |
| Roll your own tobacco | 2014 | – | – | Pictorial warnings were not mandatory | – | Only in those sections, where children's clothing and toys are sold | – | Partially | ? | 20 GEL |
|  | 2017 | – | – | ? | – | Only in those sections, where children's clothing and toys are sold | ? | Partially | 18 | 35 GEL |

| Products | Year | Sales ban | Plain packaging | Health warnings (pictorial and textual health warning) | Flavor ban | Point of sale display ban (Internal & external visibility) | Advertising ban | Smoking ban in public places<sup>11</sup> | Age limit | Taxes* (GEL) |
| --- | --- | --- | --- | --- | --- | --- | --- | --- | --- | --- |
|  | 2019 | – | – | ? | – | Only external visibility | ? | ? | 18 | 60 GEL |
|  | 2023 | – | Was adopted but not in force | ? | – | ? | ? | ? | 18 | 60 GEL |
| **Electronic nicotine delivery systems (ENDS)** | 2014 | – | – | Not specified | – | Not specified | – | Not specified | Not specified | Not specified |
|  | 2017 | – | – | Only information sheet intended for insertion into the pack | – | Only in those sections, where children's clothing and toys are sold | ? | Partially | 18 | 0.2 GEL |
|  | 2019 | – | – | Only information sheet intended for insertion into the pack | – | Only external visibility | ? | ? | 18 | 0.2 GEL |
|  | 2023 | – | – | Only information sheet intended for insertion into the pack | – | ? | ? | ? | 18 | 1.00 GEL |
| **Electronic** | 2014 | – | – | – | – | – | – | – | – | Not specified |
| non-nicotine delivery systems (ENNDS) | 2017 | – | – | – | – | – | – | Not specified | Not specified | 0.2 GEL |
|  | 2019 | – | – | – | – | – | – | Not specified | Not specified | 0.2 GEL |
|  | 2023 | – | – | – | – | ? | – | Not specified | Not specified | 1.00 GEL |
| Smokeless tobacco (chewing or snuffing tobacco) | 2014 | – | – | Pictorial warnings were not mandatory | – | Only in those sections, where children's clothing and toys are sold | – | – | ? | 20 GEL |
|  | 2017 | – | – | ? | – | Only in those sections, where children's clothing and toys are sold | ? | – | 18 | 35 GEL |
|  | 2019 | – | – | Only textual warning | – | Only external visibility | ? | – | 18 | 60 GEL |
|  | 2023 | – | – | Only textual warning | – | ? | ? | – | 18 | 60 GEL |

Notes: <sup>1</sup> In accordance with Georgian legislation, e-cigarettes and other nicotine products are classified as tobacco products. "Partially" in this column means that smoking is banned only in certain indoor places, such as educational and sports facilities, healthcare and pharmaceutical institutions, facilities storing flammable substances, and public transport.

### National Tobacco Control Policy Context

Between 2014 and 2023, Georgia strengthened its tobacco control legislation by introducing mandatory pictorial health warnings for conventional cigarettes and implementing comprehensive advertising bans (Table 3). Smoking bans in public places were well enforced, and age restrictions were standardized at 18 across all product categories.^16^ Measures such as plain packaging were enforced but with weak regulations and exempting new products such as HTPs, e-cigarettes, and no flavor bans/regulations were introduced during this period. Regulation of point-of-sale displays also advanced from limited restrictions in children’s sections to full bans of internal and external visibility by 1^st^ January of 2021.

**Table 3.** Prevalence of Current Use of Tobacco and Nicotine Products Among Adolescents in Georgia by GYTS Round and Sex.

| Year | Number of Products Used | Unweighted n | Prevalence (%) | 95% CI | Adjusted Prevalence (%) | 95% CI |
| --- | --- | --- | --- | --- | --- | --- |
| All (n=4,448; n=4,379) |  |  |  |  |  |  |
| 2014 | None | 990 | 84.7 | 81.0 – 88.4 | 85.3 | 81.8 – 88.7 |
| 2017 | None | 963 | 81.9<sup>a</sup> | 77.8 – 85.9 | 82.9 | 79.2 – 86.6 |
| 2023 | None | 1841 | 86.8<sup>a</sup> | 85.0 – 88.7 | 85.9 | 83.8 – 88.1 |
| 2014 | One product | 134 | 11.7 | 9.0 – 14.4 | 11.4 | 8.7 – 14.0 |
| 2017 | One product | 145 | 12.7 | 10.0 – 15.4 | 12.0 | 9.4 – 14.6 |
| 2023 | One product | 202 | 9.7 | 8.1 – 11.4 | 10.3 | 8.5 – 12.1 |
| 2014 | Two or more products | 38 | 3.6 | 2.2 – 5.0 | 3.4 | 2.2 – 4.5 |
| 2017 | Two or more products | 64 | 5.5 | 3.6 – 7.4 | 5.1 | 3.5 – 6.8 |
| 2023 | Two or more products | 71 | 3.5 | 2.6 – 4.4 | 3.8 | 2.7 – 4.9 |
| Boys (n=2,068; n=2,050) |  |  |  |  |  |  |
| 2014 | None | 419 | **77.4<sup>b</sup>** | 72.1 – 82.7 | 78.1<sup>i</sup> | 73.0 – 83.3 |
| 2017 | None | 425 | **75.4<sup>c</sup>** | 69.5 – 81.4 | 76.82<sup>j</sup> | 71.3 – 82.4 |
| 2023 | None | 829 | **85.6**<sup>b,c</sup> | 83.3 – 87.9 | 84.2<sup>i,j</sup> | 81.5 – 87.0 |
| 2014 | One product | 94 | **17.23**<sup>d</sup> | 13.6 – 20.9 | 16.8<sup>k</sup> | 13.0 – 20.5 |
| 2017 | One product | 85 | 15.5<sup>e</sup> | 11.7 – 19.3 | 14.7 | 11.0 – 18.4 |
| 2023 | One product | 95 | **10.1**<sup>d,e</sup> | 8.0 – 12.3 | 11.1<sup>k</sup> | 8.6 – 13.6 |
| 2014 | Two or more products | 27 | 5.4<sup>f</sup> | 3.3 – 7.5 | 5.1<sup>l</sup> | 3.3 – 6.9 |
| 2017 | Two or more products | 52 | **9.1**<sup>f,g</sup> | 6.1 – 12.1 | 8.5<sup>l,m</sup> | 5.9 – 11.1 |
| 2023 | Two or more products | 42 | **4.3**<sup>g</sup> | 3.0 – 5.5 | 4.7<sup>m</sup> | 3.2 – 6.1 |
| Girls (n=2,347; n=2,329) |  |  |  |  |  |  |
| 2014 | None | 564 | 92.1<sup>h</sup> | 89.1 – 95.1 | 92.3<sup>n</sup> | 89.4 – 95.1 |
| 2017 | None | 532 | 88.4 | 85.0 – 91.9 | 88.8 | 85.6 – 92.1 |
| 2023 | None | 1,000 | 88.1<sup>h</sup> | 85.6 – 90.5 | 87.6<sup>n</sup> | 84.7 – 90.5 |
| 2014 | One product | 38 | 6.2 | 3.6 – 8.7 | 6.1 | 3.5 – 8.6 |
| 2017 | One product | 58 | 9.6 | 6.8 – 12.4 | 9.4 | 6.8 – 11.9 |
| 2023 | One product | 104 | 9.2 | 7.3 – 11.1 | 9.4 | 7.3 – 11.5 |
| 2014 | Two or more products | 10 | 1.8 | 0.3 – 3.2 | 1.7 | 0.3 – 3.0 |
| 2017 | Two or more products | 12 | 2.0 | 0.4 – 3.5 | 1.8 | 0.3 – 3.3 |
| 2023 | Two or more products | 29 | 2.8 | 1.4 – 4.1 | 3.1 | 1.3 – 4.8 |

*Notes:* Percentages add up to 100% by year. Their totals may not sum to 100% due to rounding. Cells with unweighted n<30 should be interpreted with caution due to limited precision.
Differences in prevalence were tested using contrasts after multinomial logistic regression:
<sup>a</sup>|diff|=4.95 pps p-value=0.029; <sup>b</sup>|diff|=**8.25 pps p-value = 0.006**, <sup>c</sup>|diff|=**10.20 pps p-value = 0.002**, <sup>d</sup>|diff|=**7.10 pps p-value = 0.001**, <sup>e</sup>|diff|=5.35 pps p-value = 0.017, <sup>f</sup>|diff|=3.70 pps p-value = 0.046, <sup>g</sup>|diff|=**4.85 pps p-value = 0.004**; <sup>h</sup>|diff|=4.04 pps p-value = 0.04; <sup>i</sup>|diff|=6.11 pps p-value = 0.046; <sup>j</sup>|diff|=7.42 pps p-value = 0.021; <sup>k</sup>|diff|=5.66 pps p-value = 0.018; <sup>l</sup>|diff|=3.38 pps p-value = 0.033; <sup>m</sup>|diff|=3.82 pps p-value = 0.013; <sup>n</sup>|diff|=4.70 pps p-value = 0.029. Bolded values are statistically significant based on the Bonferroni-corrected alpha = 0.05/9=0.006.

Excise taxes on conventional cigarettes, roll-your-own tobacco, and smokeless tobacco rose steadily over the past decade, more than doubling in some cases.^21^ In contrast, new products such as e-cigarette liquids and heated tobacco were initially taxed at low levels.^21^ However, recent reforms introduced significant increases: the tax on e-cigarette liquids rose fivefold in 2023, and heated tobacco products were introduced into the tax framework in 2017 and brought under the same tax rate as conventional cigarettes, along with a 30% ad valorem tax based on retail price.^21^ Georgia’s legal framework also introduced excise stamps for tobacco and nicotine products and set import limits for personal use exemptions.^21^ Despite substantial policy progress, delays in enforcement—such as the pending implementation of plain packaging—and uneven taxation across product categories point to the need for continued efforts to reduce the affordability and appeal of new tobacco and nicotine products.

A major gap is the absence of specific regulations for non-nicotine e-cigarettes. These products are not covered under smoke-free or tobacco advertising, promotion, and sponsorship (TAPS) laws, enabling companies to market them freely and allowing their use in places where vaping or smoking is otherwise prohibited. Low, uniform taxation, regardless of whether the e-cigarette liquid contains nicotine, further increases affordability and accessibility, especially among youth, undermining the intended impact of smoke-free and TAPS policies. Moreover, regulatory gaps persist for electronic nicotine delivery devices more broadly, including limited requirements for design characteristics, packaging standards, and prominent health warnings.^16,25^

### Social Context

Between 2014 and 2023, statistically significant changes were observed across multiple social context indices related to tobacco exposure among adolescents in Georgia (Figure 1). The overall net tobacco promotion index significantly declined by 0.4 points, from –0.9 (95% CI: –1.1 to – 0.8) in 2014 to –1.3 (95% CI: –1.4 to –1.3) in 2023. Both boys and girls showed improvements, with girls experiencing a decline of 0.5 points and boys having a 0.3-point decrease, although the sex differences within each year were not statistically significant.

**Figure 1.**
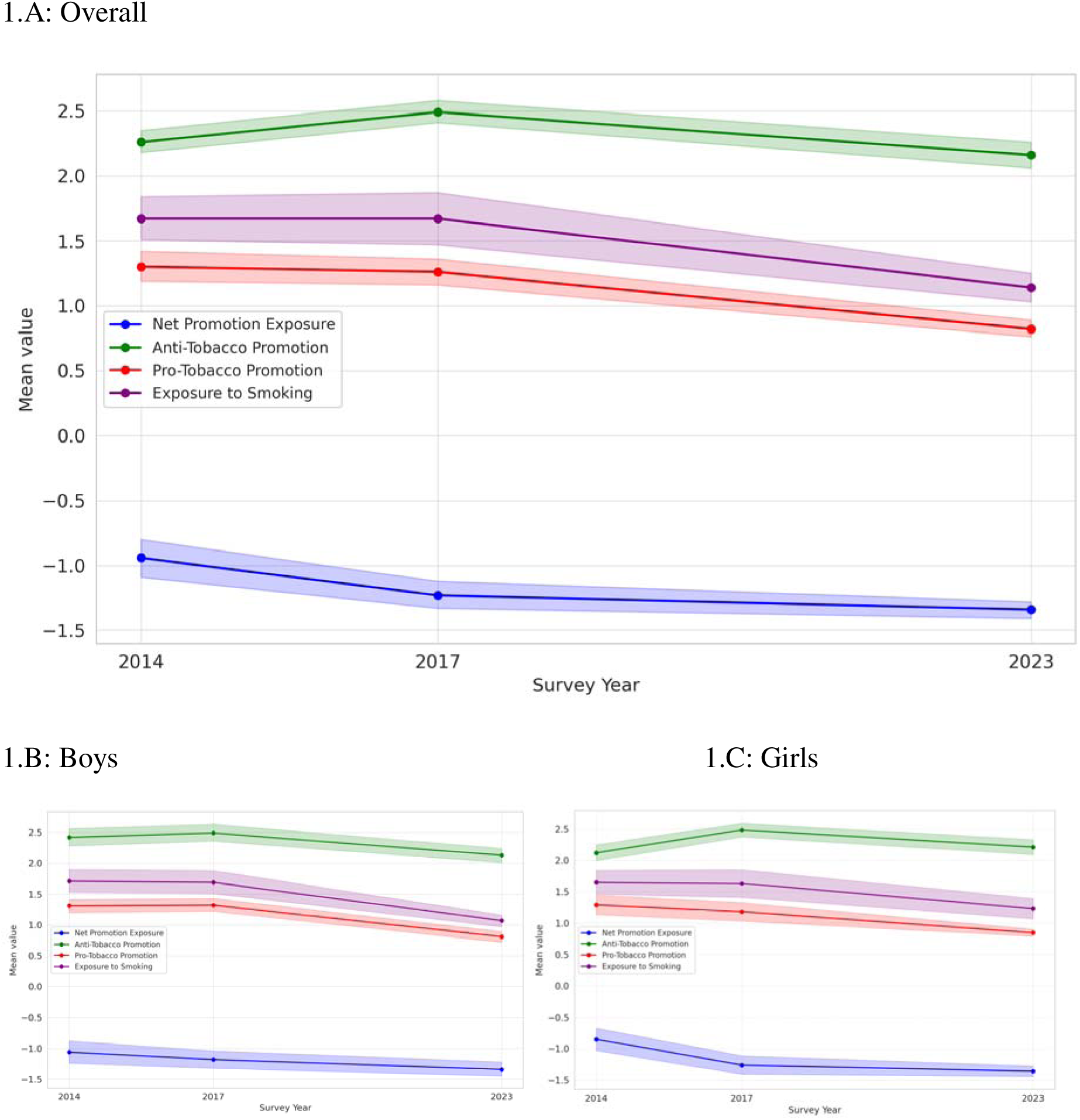
Summative Indices of Promotion Exposure to Tobacco Products among Adolescents in Georgia: Overall (A), Boys (B), and Girls (C). Note: shaded areas represent 95% CIs.

Pro-tobacco promotion exposure showed a marked and statistically significant decline of 0.5 points, from 1.3 (95% CI: 1.2–1.4) in 2014 to 0.8 (95% CI: 0.8–0.9) in 2023. The reduction was similar by sex: a 0.5-point decline among boys and 0.4 points among girls, suggesting a consistent decrease in exposure to tobacco marketing. In contrast, anti-tobacco promotion exposure increased by 0.2 points from 2.3 (95% CI: 2.2–2.4) in 2014 to a peak of 2.5 in 2017 (95% CI: 2.4–2.6) but declined by 0.3 points to 2.2 (95% CI: 2.1–2.3) in 2023. Boys experienced a 0.4-point drop between 2017 and 2023, while girls showed a 0.3-point decline during the same period.

Exposure to secondhand smoke declined significantly by 0.6 points, from 1.7 (95% CI: 1.5–1.8) in 2014 to 1.1 (95% CI: 1.0–1.3) in 2023. Boys reported a 0.6-point decrease, and girls a 0.4- point drop, although sex differences were not statistically significant.

### Trends in TNP use

Between 2014 and 2023, significant shifts occurred in current TNP use among Georgian adolescents (Table 3 and Figure 5). Among boys, the prevalence of non-use rose significantly by 8.3 pps between 2014 (77.4%) and 2023 (85.6%) (p-value = 0.006), and by 10.2 pps from 2017 (75.4%) to 2023 (p-value = 0.002). In contrast, the prevalence of non-use among girls declined from 92.1% in 2014 to 88.1% in 2023, a statistically significant 4.0-pps decrease (p-value = 0.04), although this did not meet the Bonferroni-corrected threshold for statistical significance. Among boys, both exclusive and multiple product use declined after 2017, reversing a mid-decade rise in dual or poly-use (from 5.4% in 2014 to 9.1% in 2017, before falling to 4.3% in 2023). Among girls, changes were not statistically significant, though exclusive use rose modestly from 6.2% in 2014 to around 9% in both 2017 and 2023, and dual use increased from 1.8% to 2.8% over the same period. The direction and magnitude of these trends remained consistent after adjustment for age and pocket money, with crude and adjusted estimates differing by less than one pp, suggesting minimal confounding by sociodemographic factors.

Observations with missing data for one or more product types were excluded from the ’total products used’ variable. Max number of products type used was 4. To estimate adjusted prevalence for all adolescents, age, gender, and pocket money were controlled for in the multinomial logistic regression. Gender specific adjusted prevalence was estimated controlling for age (as a continuous variable) and pocket money in the multinomial logistic regression.

Differences in prevalence were tested using contrasts after multinomial logistic regression: ^a^|diff|=4.95 pps p-value=0.029; **^b^**|**diff**|**=8.25 pps p-value = 0.006**, ^c^|**diff**|**=10.20 pps p-value = 0.002**, **^d^**|**diff**|**=7.10 pps p-value = 0.001**, ^e^|diff|=5.35 pps p-value = 0.017, ^f^|diff|=3.70 pps p-value = 0.046, **^g^**|**diff**|**=4.85 pps p-value = 0.004**; ^h^ |diff|=4.04 pps p-value = 0.04; ^i^|diff|=6.11 pps p-value = 0.046; ^j^|diff|=7.42 pps p-value = 0.021; ^k^|diff|=5.66 pps p-value = 0.018; ^l^|diff|=3.38 pps p-value = 0.033; ^m^|diff|=3.82 pps p-value = 0.013; ^n^|diff|=4.70 pps p-value = 0.029. Bolded values are statistically significant based on the Bonferroni-corrected alpha = 0.05/9=0.006.

Examining exclusive use of a single TNP offered insight into behavioral transitions not visible in product-group prevalence. This analysis revealed pronounced sex-specific shifts in product preferences. Among adolescents who reported using only one type of TNP, the distribution of products (e.g., cigarettes vs. e-cigarettes vs. other products) changed significantly over time, with notable differences between boys and girls (Figure 2). This analysis focused on exclusive use of a single TNP provides additional insight into behavioral transitions that are not evident from aggregate prevalence estimates alone.

**Figure 2.**
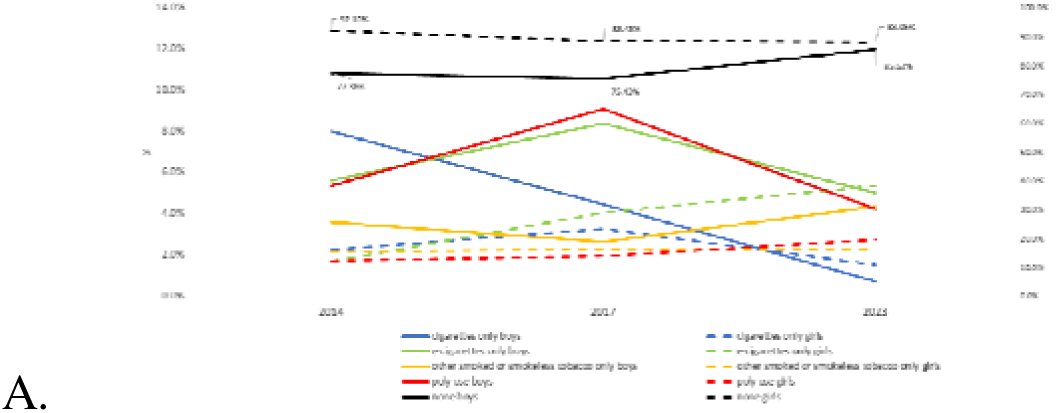
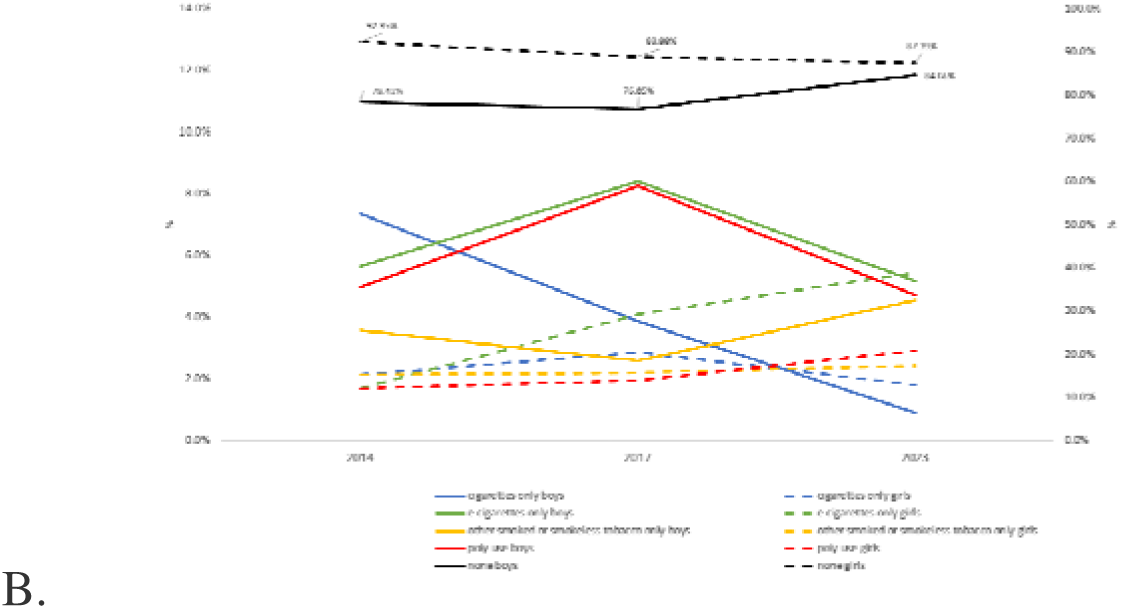
(A: Unadjusted; B: Adjusted) Prevalence of TNP Use by Type and Number Among Adolescents in Georgia, GYTS 2014, 2017, and 2023. *Note:* In Figure 1B, prevalence was adjusted for students’ age and pocket money.

Exclusive cigarette use declined for both sexes but more steeply among boys, with a 36.8 pps drop compared to a 13.1 pps decline among girls (p-value = 0.035). In contrast, exclusive e-cigarette use rose among girls throughout the period, increasing by 26.2 pps, while among boys it rose initially and then declined slightly, resulting in a smaller net increase of 16.2 pps. By 2023, girls had a significantly higher adjusted prevalence of exclusive e-cigarette use than boys.

Additionally, exclusive use of other smoked or smokeless tobacco products increased by 20.6 pps among boys but decreased by 13.1 pps among girls, resulting in a significant widening of the sex gap by 33.7 pps (p-value = 0.005). These shifts underscore a growing divergence in product preference and use patterns between girls and boys, particularly in relation to new and non-cigarette products.

## CONCLUSION

Between 2014 and 2023, the use of TNPs among adolescents in Georgia underwent important shifts, shaped by evolving product landscapes, marketing strategies, and regulatory measures. Overall, the prevalence of non-use remained relatively high, hovering around 85%, but patterns differed by sex and product type. Boys experienced statistically significant reductions in both single and multiple product use. These positive trends coincide with major national tobacco control measures implemented since 2017, including the comprehensive tobacco law enacted in 2018 and substantial increases in excise taxes. These findings reaffirm prior evidence that legislative interventions, including higher age-of-sale restrictions, increased taxation, and stronger advertising and smoke-free protections, are effective in reducing youth smoking. Georgia implemented each of these measures during 2014-2023.^26^

However, product use patterns among girls followed a different trajectory. While overall TNP use among girls remained lower than among boys, exclusive e-cigarette use rose steadily over the study period. By 2023, girls had surpassed boys in exclusive e-cigarette use—a reversal of historical gender patterns observed in Georgia and noted in a few international studies that specifically examined exclusive use.^27-29^ Evidence from other contexts suggests that girls may be more likely to initiate e-cigarette use, perceive them as less harmful, and be influenced by targeted marketing and social media trends, particularly those leveraging flavours, sleek designs, and gendered messaging.^28-29^

The social environment in which youth engage with tobacco-related promotion evolved significantly. From 2014 to 2023, exposure to pro-tobacco advertising and promotion and secondhand smoke declined steadily, likely contributing to reduced poly-use and greater resistance to social pro-tobacco cues, especially among boys. The net promotion exposure index showed improvements across all groups, suggesting a broad shift toward less permissive social norms. However, anti-tobacco promotion exposure peaked in 2017 and declined by 2023, potentially signaling reduced investment in school-based education and public awareness efforts, a trend also seen in broader European contexts.^30^ In Georgia, fewer adolescents reported learning about tobacco harms in school in 2023 than in previous rounds, a concerning development that could compromise future progress.

Sex-stratified analyses further highlighted divergent trends in psychosocial exposures. Although both boys and girls showed declines in pro-tobacco advertising and promotion exposure and secondhand smoke, improvements in the net promotion index were slightly less pronounced among girls. This suggests that girls may be facing more subtle or indirect forms of tobacco marketing, possibly through social media or peer networks. In parallel, exclusive use of e-cigarettes and smokeless tobacco products rose more sharply among girls – patterns that require tailored public health interventions.

Exclusive use patterns reinforced gender differences. Among boys, poly-use was most prevalent in 2017, followed by a significant decline by 2023. Among girls, e-cigarettes became the dominant product among those reporting exclusive use. These findings align with global reports indicating that TNP marketing is increasingly tailored to adolescents’ gender identity and lifestyle preferences.^31,32^

Despite recent policy wins in Georgia, including excise tax hikes and a national ban on indoor smoking, important regulatory gaps remain. E-cigarettes, nicotine pouches, and other new products are not yet subject to comprehensive packaging, flavor restrictions, or design restrictions. Enforcement of marketing bans, particularly on digital platforms, remains weak. Temporary tax exemptions (e.g., on raw tobacco in 2024) and aggressive industry lobbying further dilute policy effectiveness. Alarmingly, the tobacco industry continues to co-opt healthcare professionals to promote “harm reduction” narratives – tactics that must be actively countered.

### Strengths and Limitations

Our study draws on nationally representative, school-based survey data with strong response rates and standardized methodologies across three time points. The use of social context indices added depth to the analysis, enabling us to move beyond simple sex-disaggregation of prevalence to consider how contextual factors shifted over time. This approach responds to recent calls for gender responsive tobacco control research, which emphasize that while sex disaggregated analyses provide important baseline insights, they often fail to capture the nuanced ways gender norms, social positioning, and targeted marketing intersect to shape tobacco use.^7^ While our approach is not a complete solution, it represents an important step toward more comprehensive gender responsive monitoring, and provides a foundation for further refinement and expansion in future research.

Like other GYTS analyses, our study is subject to certain common limitations, including reliance on self-reported data and the cross-sectional design, which precludes causal inference. In Georgia’s context, the most relevant additional consideration is that school-based sampling may underrepresent adolescents who are not enrolled or frequently absent; however, secondary school enrollment is nearly universal, making this limitation minimal.^33^ Additionally, some subgroup estimates (e.g., Table 3) were based on small cell sizes and should be interpreted with caution.

In conclusion, Georgia’s tobacco control efforts have achieved substantial progress over the past decade, particularly in reducing conventional tobacco use among adolescent boys and improving the social environment around tobacco. However, rising e-cigarette use among girls, persistent gaps in regulation and enforcement, and the rapid proliferation of new TNPs pose serious challenges. These patterns reflect broader global concerns that public health responses to tobacco remain largely gender-blind, in contrast to industry marketing that has long manipulated gender norms to expand its consumer base. As highlighted by the WHO’s 2018 policy brief on gender-responsive tobacco control, meaningful progress requires interventions that explicitly address how gender influences product appeal, access, and exposure.^8^ Addressing these issues in Georgia will require recalibrated, evidence-based, and gender-sensitive strategies, backed by strong surveillance, consistent enforcement, public education, and insulation from industry interference, to safeguard the next generation from tobacco-related harm.

## Data Availability

All data produced in the present work are contained in the manuscript

## Ethics statements

Patient consent for publication: Not applicable.

Ethics approval: The GYTS data are de-identified and publicly available; hence, its analysis does not constitute human subjects’ research. The study was exempt from review by the research ethics boards at the investigators’ institutions.

Disclaimer: Funding was provided by the Bloomberg Initiative to Reduce Tobacco Use through the CDC Foundation with a grant from <u>Bloomberg Philanthropies</u>. The findings and conclusions in this publication are those of the authors and do not necessarily represent the official position of the CDC Foundation.

## Contributions

AC, KG, and YT conceptualized the study. KG, GB, VG, and YT conducted quantitative analyses. AD, GB, and TA carried out qualitative analyses, including review of legislation in Georgia. AD, GB, KG, TA, and YT drafted the manuscript. AC and LS provided critical revisions. All authors read and approved the final manuscript.

## Funding statement

The authors gratefully acknowledge funding provided by the Bloomberg Initiative to Reduce Tobacco Use through the CDC Foundation with a grant from *<u>Bloomberg Philanthropies</u>.* The findings and conclusions in this publication are those of the authors and do not necessarily represent the official position of the CDC Foundation.

## Competing interests

None declared.

## Patient and public involvement

Patients and/or the public were not involved in the design, or conduct, or reporting or dissemination plans of this research based of secondary data analysis.

---

**Supplemental Table 1.** Summative Indices of Promotion Exposure to Tobacco Products among Adolescents in Georgia.

| Year | Study population | Unweighted n | Unadjusted mean (%) | 95% CI |
| --- | --- | --- | --- | --- |
| Net promotion exposure<sup>1</sup> |  |  |  |  |
| 2014 | All | 1,094 | -0.94 | -1.09 – -0.80 |
| 2017 | All | 974 | -1.23 | -1.33 – -1.12 |
| **2023** | **All** | **1,580** | **-1.34** | **-1.41 – -1.28** |
| 2014 | Boys | 488 | -1.06 | -1.24 – -0.88 |
| 2017 | Boys | 450 | -1.18 | -1.32 – -1.04 |
| **2023** | **Boys** | **741** | **-1.34** | **-1.45 – -1.22** |
| 2014 | Girls | 595 | -0.85 | -1.03 – -0.67 |
| 2017 | Girls | 518 | -1.26 | -1.41 – -1.12 |
| **2023** | **Girls** | **831** | **-1.36** | **-1.45 – -1.28** |
| Anti-tobacco promotion exposure (range 0/lowest – 6/highest)<sup>2</sup> |  |  |  |  |
| 2014 | All | 1,149 | 2.26 | 2.18 – 2.35 |
| 2017 | All | 1,073 | 2.49 | 2.41 – 2.58 |
| 2023 | All | 1,784 | 2.16 | 2.06 – 2.26 |
| 2014 | Boys | 527 | 2.42 | 2.28 – 2.56 |
| 2017 | Boys | 508 | 2.49 | 2.36 – 2.63 |
| 2023 | Boys | 832 | 2.13 | 2.01 – 2.24 |
| 2014 | Girls | 611 | 2.12 | 2.00 – 2.25 |
| 2017 | Girls | 558 | 2.48 | 2.37 – 2.59 |
| 2023 | Girls | 939 | 2.21 | 2.09 – 2.32 |
| Pro-tobacco promotion exposure (range 0/lowest – 3-4/highest)<sup>3</sup> |  |  |  |  |
| 2014 | All | 1,257 | 1.30 | 1.19 – 1.42 |
| 2017 | All | 1,165 | 1.26 | 1.16 – 1.36 |
| **2023** | **All** | **2,092** | **0.82** | **0.76 – 0.89** |
| 2014 | Boys | 584 | 1.31 | 1.20 – 1.41 |
| 2017 | Boys | 544 | 1.32 | 1.22 – 1.43 |
| **2023** | **Boys** | **972** | **0.81** | **0.72 – 0.89** |
| 2014 | Girls | 657 | 1.29 | 1.14 – 1.45 |
| 2017 | Girls | 613 | 1.18 | 1.04 – 1.33 |
| **2023** | **Girls** | **1,103** | **0.85** | **0.79 – 0.90** |
| Exposure to smoking<sup>4</sup> (range 0/lowest to 6/highest) |  |  |  |  |
| 2014 | All | 1,086 | 1.67 | 1.51 – 1.84 |
| 2017 | All | 1,067 | 1.67 | 1.47 – 1.87 |
| **2023** | **All** | **2,227** | **1.14** | **1.03 – 1.25** |
| 2014 | Boys | 504 | 1.71 | 1.53 – 1.90 |
| 2017 | Boys | 513 | 1.69 | 1.51 – 1.88 |
| **2023** | **Boys** | **1,056** | **1.07** | **0.97 – 1.16** |
| 2014 | Girls | 569 | 1.65 | 1.46 – 1.84 |
| 2017 | Girls | 547 | 1.63 | 1.41 – 1.85 |
| **2023** | **Girls** | **1,154** | **1.23** | **1.07 – 1.39** |

## Notes

### Competing Interest Statement

The authors have declared no competing interest.

### Funding Statement

Funding was provided by the Bloomberg Initiative to Reduce Tobacco Use through the CDC Foundation with a grant from Bloomberg Philanthropies. The findings and conclusions in this publication are those of the authors and do not necessarily represent the official position of the CDC Foundation

### Author Declarations

The GYTS data are de-identified and publicly available; hence, its analysis does not constitute human subjects research

